# Generic Patient-Reported Outcomes and Outcome Measures: A Systematic Review Study Protocol

**DOI:** 10.1101/2021.05.28.21257980

**Authors:** Yuki Seidler, Erika Mosor, Margaret Andrews, Carolina Watson, Nick Bott, Evelyn Gross, Jan A Hazelzet, Maisa Omara, Valentin Ritschl, Tanja Stamm

## Abstract

**Background:** Patient-reported outcomes (PROs) are an essential part of health outcome measurement and vital to patient-centricity and valued-based care. Several international consortia have developed core outcome sets and many of them include PROs. PROs are measured by patient-reported outcome measures (PROMs). PROs and PROMs can be generic or specific to certain diseases or conditions. While the characteristics of generic PROs and PROMs are well recognised as widely relevant and applicable across different domains, diseases and conditions, there is a lack of knowledge on the types of PROs measured by generic PROMs. We also do not know in which disease areas generic PROs and PROMs are commonly used. To date, there has been no systematic review solely focusing on generic PROMs, what they measure and their areas of application.

**Objectives:** This systematic review will identify core PROs measured by generic PROMs used in adult populations and the areas in which they are applied.

**Methods:** We will conduct a systematic review of reviews. The screening process and the reporting will comply with the PRISMA (Preferred Reporting Items for Systematic Reviews and Meta-Analysis) 2020 Statement. We will use four databases, Medline [PubMed], CINHAL [Ebsco], Cochrane [Cochrane Library], and PsycINFO [Ovid], and reports from international consortia. Inclusion criteria are systematic reviews, meta-analysis or patient-reported outcome sets developed by international consortia reporting on generic PROMs in adult populations. Articles primarily focusing on patient-reported experience measures (PREMs), children or adolescents, or those not written in English will be excluded. Risk of bias will be assessed by checking if the included articles comply with established guidelines for systematic reviews such as the PRISMA statement. We will extract generic PROMs and PROs measured by these PROMs, and the areas applied from the selected articles and reports. Extracted data and information will be quantitatively and qualitatively synthesised without statistical interference. The quality of the synthesised evidences will be assessed by clarifying the strengths, limitations and possible biases in our review.

## Introduction

Patient-reported outcomes (PROs) are an essential part of health outcome measurement and vital to patient-centricity and valued-based care. Several international consortia have developed core outcome sets and many of them include PROs. PROs are measured by patient-reported outcome measures (PROMs). PROs and PROMs can be generic or disease- or condition-specific. Disease- or condition-specific PROMs are suitable for measuring outcomes of a single disease or a specific condition [1-3]. Generic PROs and PROMs are neither disease nor condition specific. They capture the broad array of patient experiences with an initial focus on the health-related quality of life [4] and cover the broad “spectrum of function, disability and distress that is relevant to quality of life” [5]. They should be applicable to all sexes, different types and severities of diseases, and different treatment types and interventions. Ideally, generic PROMs should be used in different cultural settings and ethnic populations, too [4]. In this review, PROs and PROMs are considered generic when they are “applicable (and relevant) to a broad range of patient groups, diseases, and interventions” [6] and measure at least one or more of the general aspects of health or wellbeing as defined by the World Health Organisation — physical, mental or social health [7].

Several international consortia have developed core outcome sets that include both generic and disease- or condition specific PROs and PROMs. Examples of such initiatives are the Core Outcome Set/COMET (www.comet-initiative.org), the Outcome Measures Framework/OMF (https://effectivehealthcare.ahrq.gov), the International Classification of Functioning, Disability and Health (ICF) Core Sets Projects (www.icf-research-branch.org), the International Consortium for Health Outcome Measurements/ICHOM (www.ichom.org) and the Patient-Reported Outcomes Measurement Information System/PROMIS (www.healthmeasures.net). These initiatives have different foci and aims. COMET and OMF developed core sets of outcomes; COMET ones for specific conditions [8] and OMF such that are applicable across conditions [9]. ICF-oriented researchers established domain sets in the area of functioning and health without suggesting concrete instruments [10]. ICHOM and PROMIS proposed both PROs and PROMs. One of the crucial goals of PROMIS, for example, is to develop a set of “universally-relevant measures” (generic PROMs) that are widely relevant and applicable to chronic disorders and diseases [11].

Despite these recognised characteristics of generic PROs and PROMs and related international initiatives, there is a lack of synthesised knowledge on the types of PROs measured by generic PROMs, and in which context or diseases areas they are used. There has been no systematic review solely focusing on generic PROMs and their areas of application to date.

### Objectives

This systematic review will identify core PROs measured by generic PROMs used in adult populations including the medical areas in which they are applied. Textbox 1 shows the population, intervention, comparison, and outcome (PICO) of the planned review.

#### Textbox 1. Questions addressed in the review using PICO

|  |  |
| --- | --- |
| **Population** | All adults (patients and non-patients) |
| **Intervention** | Generic patient-reported outcome measures (PROMs) |
| **Comparison** | Not applicable |
| **Outcome** | Identification of PROs measured by generic PROMs and areas of application |

## Methods

This protocol was developed using the PRISMA-P (Preferred Reporting Items for Systematic Reviews and Meta-Analysis Protocols) 2015 Statement as a guideline [12]. The checklist is attached as a supplemental document. The review will be conducted in compliance with the PRISMA 2020 Statement: an updated guideline for reporting systematic reviews [13].

### Eligibility criteria

Systematic reviews, meta-analysis and reports published by international consortia will be included. Detailed inclusion and exclusion criteria are summarised in Textbox 2. We will limit our database search to systematic reviews because of the large number of published articles on generic PROMs and to build on existing knowledge and to integrate insights regarding generic PROs and PROMs. For example, a quick scanning search conducted in January 2021 resulted in more than 20,000 articles containing the word “SF-36 (Short Form Health Surveys, 36 items)”, one of the most commonly applied generic PROMs, in the abstract or title field of the database Medline [PubMed].

#### Textbox 2. Inclusion and exclusion criteria

| **Include** | **Exclude** |
| --- | --- |
| • Systematic reviews, meta-analysis or reports on outcome sets developed by international consortia containing generic PROMs in adult populations.<br>• Articles from all countries and geographical locations. | • Articles with a primary focus on:<br>▪ Children or adolescents (aged <19)<br>▪ Dissertations, guideline statements, and abstracts<br>▪ Non-English articles |

### Context

The review context will encompass all healthcare settings, including acute, outpatient, primary, and community care. We will include systematic reviews containing patient data from trial registries, databases, and patient groups.

### Information sources

Medline [PubMed], CINHAL [Ebsco], Cochrane [Cochrane library] PsycINFO [Ovid] will be used to search systematic reviews or meta-analysis that meet the eligibility criteria. Patient-reported outcome sets published by international consortia, and missing articles identified from the screening of the reference lists of the extracted articles will be added to the review. No restriction on the year of publication will be applied.

### Search Strategies

Examples of preliminary search strategies for Medline[PubMed] include: (“Patient Reported Outcome Measures”[MeSH Terms] OR “patient reported outcome*”[Title/Abstract]) AND “systematic review”[Filter]. (“PROMIS”[Title/Abstract] OR “patient reported outcomes measurement information system”[Title/Abstract]) AND “systematic review”[Filter]. “International Consortium for Health Outcomes Measurement” [All Fields].

### Data management

All identified literature will be imported into Endnote X9^™^ (Thomson Reuters, Philadelphia, PA, USA), and duplicates will be removed. Documentation of search results will be saved in the institute’s internal secure server hosted by the Medical University of Vienna.

### Selection process

First, two reviewers, YS and VR, will screen the titles and abstracts of all retrieved articles and then screen the full texts of the selected articles according to the inclusion and exclusion criteria. Any discrepancies in the results or disagreements will be discussed and resolved until consensus is met with a third reviewer (TS). The reference list of the selected article will be screened and considered for inclusion. The screening and selection processes will be recorded using the PRISMA-P flow diagram. The numbers of excluded literature and the reasons will be stated in a diagram and a table.

### Data collection process and data items

Data will be extracted by the team members (MA, CWB, EG, EM, MO, YS and VR). Clear instructions, templates and examples will be given in written on the types of data to be extracted. These include publication year, study designs, the purpose of the review, generic PROMs, PROs measured by generic PROMs, and disease areas or context in which the generic PROMs are applied. When necessary, the original studies will be screened to extract essential data.

### Outcomes and prioritisation

The outcomes of this systematic review are generic PROMs that have been used widely across diseases, types of PROs measured by these generic PROMs and their areas of applications.

### Data synthesis

Our review does not assess the strength or the effects of particular interventions. Therefore, extracted data and information will be quantitatively and qualitatively synthesised without statistical interference and will be visualised in tables and figures.

### Meta-biases and risk of bias assessment

Articles to be included in the review will go through a critical appraisal process by two reviewers (YS and VR) supervised by the methodologist (TS). For example, the reviewers will check if the reviews or meta-analysis comply with established guidelines such as the PRISMA statement or if outcome sets developed by international consortia have followed a rigorous literature review and expert consensus processes. The study quality and risk bias of the original studies will be directly obtained from the published systematic reviews or meta-analysis.

### Ensuring confidence in the strength of the synthesised evidences

The quality of the synthesised evidences will be assessed by clarifying the strengths, limitations and possible biases in our review.

### Contributions of protocol authors

YS and EM drafted the protocol supervised by TS. VR and YS developed the search strategies. All authors read and commented on the draft protocol and approved the final version. TS is the guarantor of the review.

## Data Availability

The study is literature review based on a secondary data analysis.

